# Influence of vitamin D supplementation on fracture risk, bone mineral density and bone biochemistry in Mongolian schoolchildren: multicenter double-blind randomized placebo-controlled trial

**DOI:** 10.1101/2023.05.18.23290181

**Authors:** Davaasambuu Ganmaa, Polyna Khudyakov, Uyanga Buyanjargal, Enkhtsetseg Tserenkhuu, Sumiya Erdenenbaatar, Chuluun-Erdene Achtai, Narankhuu Yansan, Baigal Delgererekh, Munkhzaya Ankhbat, Enkhjargal Tsendjav, Batbayar Ochirbat, Badamtsetseg Jargalsaikhan, Enkhmaa Davaasambuu, Adrian R Martineau

**Affiliations:** Channing Division of Network Medicine, Brigham and Women’s Hospital, Harvard Medical School, Boston, MA 02115, USA; Harvard T.H. Chan School of Public Health, Boston, MA 02115, USA; Sage Therapeutics, Cambridge, MA, USA; Mongolian Health Initiative, Royal Plaza, Bayanzurkh District, Ulaanbaatar 13312, Mongolia; Global Laboratory, Royal Plaza, Bayanzurkh District, Ulaanbaatar 13312, Mongolia; National Center for Communicable Disease, Bayanzurkh District, Ulaanbaatar 13312, Mongolia; Mongolian National University of Medical Sciences, Sukhbaatar District, Ulaanbaatar 14210, Mongolia; National Center for Maternal and Child Health, Bayangol District, Ulaanbaatar 16060, Mongolia; Blizard Institute, Barts and The London School of Medicine and Dentistry, Queen Mary University of London, London E1 2AT, UK

## Abstract

**Background:** Randomized controlled trials (RCT) of vitamin D supplementation to reduce fracture risk in children are lacking.

**Methods:** We conducted a Phase 3 RCT of weekly oral supplementation with 14,000 IU vitamin D_3_ for 3 years in Mongolian schoolchildren aged 6-13 years. Serum 25-hydroxyvitamin D (25[OH]D) concentrations and the proportion of participants reporting ≥1 fracture were secondary outcomes for the main trial. Radial bone mineral density (BMD) was assessed in a nested sub-study, with serum concentrations of parathyroid hormone (PTH) and bone-specific alkaline phosphatase (BALP) determined in a subset of participants.

**Findings:** 8851 children were enrolled in the main trial, of whom 1465 also participated in the sub-study. Vitamin D deficiency was prevalent at baseline (25[OH]D <20 ng/mL in 90.1%). The intervention elevated 25(OH)D concentrations (adjusted inter-arm mean difference [aMD] 20.3 ng/mL, 95% CI 19.9 to 20.6) and suppressed PTH concentrations (aMD −13.6 pmol/L, 95% CI −23.5 to −3.7), but it did not influence fracture risk (adjusted risk ratio 1.10, 95% CI 0.93 to 1.29, P=0.27) or radial BMD z-score (aMD −0.06, 95% CI −0.18 to 0.07, P=0.36). Vitamin D suppressed serum BALP concentrations more among participants with baseline 25(OH)D concentrations <10 vs. ≥10 ng/mL (P_interaction_=0.04). However, effects of the intervention on fracture risk and radial BMD were not modified by baseline vitamin D status (P_interaction_≥0.67).

**Interpretation:** Weekly oral vitamin D supplementation elevated serum 25(OH)D concentrations and suppressed PTH concentrations in vitamin D-deficient schoolchildren in Mongolia. However, this was not associated with reduced fracture risk or increased radial BMD.

**Funding:** National Institutes of Health

**RESEARCH IN CONTEXT:** *EVIDENCE BEFORE THIS STUDY:* We searched PubMed from inception to 31^st^ December 2022 for randomized controlled trials (RCT) evaluating effects of vitamin D supplementation on bone mineral content (BMC), bone mineral density (BMD) and fracture risk in HIV-uninfected schoolchildren. A meta-analysis of data from 884 participants in six RCT reported no statistically significant effects of vitamin D on total body BMC, hip BMD, or forearm BMD, but a trend towards a small positive effect on lumbar spine BMD. RCT investigating fracture outcomes were lacking, as were RCT investigating effects of vitamin D on bone outcomes in children with baseline serum 25-hydroxyvitamin D (25[OH]D) concentrations <20 ng/mL.

*ADDED VALUE OF THIS STUDY:* This is the first RCT to investigate effects of vitamin D supplementation on fracture risk and BMD in Mongolian schoolchildren. Vitamin D deficiency was prevalent among the study population at baseline, and weekly oral supplementation with 14,000 IU vitamin D_3_ for 3 years elevated serum 25(OH)D concentrations into the physiologic range and suppressed serum PTH concentrations. However, the intervention did not influence fracture risk or radial BMD, either in the study population as a whole or in the large sub-group of participants with baseline serum 25(OH)D concentrations <10 ng/mL.

*IMPLICATIONS OF ALL THE AVAILABLE EVIDENCE:* Taken together with null findings from another recenty-completed phase 3 RCT of weekly oral vitamin D supplementation conducted in South African schoolchildren, our findings do not support a role for vitamin D supplementation to reduce fracture risk or increase BMD in primary schoolchildren.

## INTRODUCTION

Around a third of children sustain at least one fracture before the age of 18 years, with the risk peaking around the time of the pubertal growth spurt.^1,2^ Impacts range from temporary limitation of activity through hospitalization and occasionally permanent disability.^3^ Injury prevention represents one strategy to address this problem,^4^ but a complementary approach relates to development of interventions to improve bone strength in childhood.^5^ The potential for vitamin D supplementation to achieve this end has attracted considerable attention, based on the physiologic role of its active metabolite calcitriol in supporting calcium absorption^6^ together with findings from observational studies reporting a high prevalence of vitamin D deficiency among children in diverse settings, associating independently with increased risk of fractures.^7^ However, randomized controlled trials (RCT) of vitamin D supplementation to reduce risk of fractures in children are lacking.

An opportunity to address this evidence gap arose when we conducted a Phase 3 RCT of weekly oral vitamin D supplementation for 3 years in 8,851 school children aged 6-13 years living in Mongolia, a setting with a particularly high fracture burden^8^ where vitamin D deficiency is also widely prevalent.^9^ The primary outcome for the trial was incidence of tuberculosis infection;^10^ secondary outcomes included fracture incidence and serum concentrations of 25-hydroxyvitamin D (25[OH]D) in all participants, and bone mineral density in a subset of 1,465 participants who took part in a nested bone mineral density (BMD) sub-study, in which radial quantitative ultrasound (QUS) was performed at baseline and annual intervals thereafter. A subset of 100 BMD sub-study participants also contributed serum for determination of concentrations of parathyroid hormone (PTH), calcium, albumin, total alkaline phosphatase (ALP) and bone-specific alkaline phosphatase (BALP) at baseline and 3-year follow-up.

## METHODS

### Study Design, Setting, Participants

We conducted a parallel two-arm double-blind individually randomized placebo-controlled trial in eighteen public schools in Ulaanbaatar, Mongolia, as previously described.^10,11^ The primary outcome of the trial was acquisition of latent tuberculosis infection; the current manuscript reports effects of the intervention on fracture risk, bone mineral density and bone biochemistry outcomes. Principal inclusion criteria were age 6-13 years at screening and attendance at a participating school; principal exclusion criteria were a positive QuantiFERON-TB Gold in-tube assay (QFT) result, presence of conditions associated with vitamin D hypersensitivity (primary hyperparathyroidism or sarcoidosis) or immunocompromise (taking immunosuppressant medication or cytotoxic therapy), use of vitamin D supplements, signs of rickets (all participants were screened for rickets via physical examination by a pediatrician), or intention to move from Ulaanbaatar within 4 years of enrolment.

### Randomization and masking

Eligible participants were individually randomized to receive a weekly capsule containing 14,000 IU (350 μg) vitamin D_3_ or placebo for three years, with a one-to-one allocation ratio and stratification by school of attendance. Treatment allocation was concealed from participants, care providers, and all trial staff (including senior investigators and those assessing outcomes) so that the double-blind was maintained. Further details of randomization and masking procedures are provided in Supplementary Material.

### Ethics and Regulatory

Each child and their parent/guardian provided written informed assent/consent prior to participation. The study was approved by institutional review boards of the Mongolian Ministry of Health, Mongolian National University, and Harvard T. H. Chan School of Public Health (IRB # 14-0513).

### Baseline Procedures

At baseline, participants’ parents were asked to complete questionnaires detailing their socioeconomic circumstances, lifestyle and dietary factors influencing vitamin D status, and intake of foods previously shown to be major contributors to dietary calcium intake in urban Mongolia.^12^ Participating children’s height was then measured using a portable stadiometer (SECA, Hamburg, Germany), and their weight was measured using a Digital Floor Scale (SECA). In a subset of 1,465 participants attending one of fourteen participating schools, randomly selected with stratification by school year to ensure equal representation from 2^nd^ to 6^th^ year students, BMD and speed-of-sound were measured at the distal one third of the radius using a Sunlight MiniOmni™ portable bone sonometer (BeamMed, Petah Tikva, Israel) according to the manufacturer’s instructions.^13^ Radial BMD was measured on the non-dominant arm, unless this was injured, in which case measurement was done on the dominant arm.

Finally, a blood sample was drawn for QFT testing and separation and storage of serum for determination of 25(OH)D concentrations (all participants) and calcium, albumin, PTH, total ALP and BALP (randomly selected n=100 subset of BMD sub-study participants).

### Follow-Up Procedures

During school terms, study participants had weekly face-to-face visits at which study medication was administered and adverse events were recorded. During school holidays, children were either given a single bolus dose of up to 36,000 IU (shorter holidays), study staff travelled to participants homes to administer medication, or parents were supplied with sufficient trial medication to cover the holiday period, along with instructions on its storage and administration. At 2- and 3-year follow-up, history of fractures was captured using an electronic case report form (Fig. S1, Supplementary Material). Repeat quantitative radial ultrasound was performed in BMD sub-study participants at 1-, 2- and 3-year follow-up, using the same method as at baseline. At 3-year follow-up, a second blood sample was drawn from all participants for QFT testing and separation and storage of serum for determination of, 25(OH)D concentrations (all participants) and calcium, albumin, PTH, total ALP and BALP (n=100 subset for whom these parameters were also measured at baseline).

### Measurement of vitamin D status

25(OH)D concentrations were determined in serum samples from baseline and 3-year follow-up using an enzyme-linked fluorescent assay (VIDAS 25OH Vitamin D total, bioMérieux, Marcy-l’Étoile, France). These measurements were validated and standardized using standards provided by the Vitamin D External Quality Assessment Scheme [23]. Total CV was 7.9%, mean bias was 7.7% and the limit of quantitation (LOQ) was 8.1 ng/ml. Non-zero 25(OH)D values were standardized using a published method [24], utilizing a set of 40 serum samples provided by DEQAS (the Vitamin D External Quality Assessment Scheme, http://www.deqas.org/). Values below the LOQ were classified as <8.1 ng/ml.

### Measurement of serum concentrations of bone turnover markers

Serum concentrations of PTH, calcium, albumin and total ALP were determined using a COBAS e 411 analyzer (Roche Diagnostics, Indianapolis, IN, USA) in the ISO 15189-accredited integrated clinical laboratory at the National Second Central Hospital, Ulaanbaatar, Mongolia. Albumin-adjusted calcium (mmol/L) was calculated as total calcium (mmol/L) + 0.02 × (40 – albumin [g/L]). Serum concentrations of BALP were determined at the Global Laboratory, Ulaanbaatar, Mongolia, using MicroVue bone alkaline phosphatase enzyme immunoassay kits (Quidel Corporation, San Diego, CA, USA) performed according to the manufacturer’s instructions.

### Outcomes

The primary bone outcome of interest was the proportion of participants in the main trial reporting one or more fractures during follow-up. Secondary outcomes were radial BMD and speed-of-sound (all BMD sub-study participants) and serum concentrations of 25(OH)D, PTH, adjusted calcium, total ALP and BALP (n=100 subset of BMD sub-study participants).

### Sample Size and Statistical Methods

The sample size calculation for the main trial was based on the power needed to detect a clinically significant effect of the intervention on the primary endpoint (incident latent tuberculosis infection) as described previously.^1^ For the BMD sub-study, we calculated that a total of 1,464 participants (732 per arm) would need to be recruited to demonstrate a difference of 0.2 z-scores between arms with alpha = 0.05 and power = 80%, assuming standard deviation for BMD z-score of 1.1 and 35% loss to follow-up. We originally planned to analyze bone biochemistry parameters in all BMD sub-study participants, but resource constraints limited these analyses to a n=100 subset.

Estimated calcium intakes were calculated as previously described^11^ on the basis of parental responses to a one-off food frequency questionnaire administered in March 2018, which captured participants’ frequency of intake of calcium-containing foods, and the calcium content of those foods, based on food composition data compiled from analysis of their calcium content. Serum 25(OH)D values were adjusted for seasonal variation prior to analysis using a sinusoidal model, as previously described.^4^ BMI was calculated as weight (kg)/ (height [m])^2^. Fracture data were cleaned to reduce risk of double-reporting: where two or more fractures were reported to occur at the same anatomical site within 28 days of each other by the same participants, they were adjudged as likely duplicates of the same event, and only one instance was retained. The proportion of participants reporting one or more fractures was analyzed using generalized linear models with binomial distribution and a log link function with adjustment for school of attendance; treatment effects were presented as adjusted risk ratios with 95% confidence intervals. Radial BMD z-scores and speed-of-sound measurements were analyzed overall and in each sub-group using mixed models for repeated measures (MMRM) with random effects for individuals and mixed effects for treatment, time and treatment-by-time interaction, adjusted for school of attendance. Least-squares (LS) means with 95% confidence intervals (CI) were reported for treatment differences at each annual follow-up time point. Statistical significance of treatment effects was reported for each follow-up time point and for the overall treatment-by-time interaction. Significance of sub-group effects was calculated using separate MMRM models as described above but with additional inclusion of a treatment-by-subgroup interaction term. Biochemical parameters were analyzed using linear regression with adjustment for baseline values and school of attendance; LS means for inter-group differences were presented with 95% CI. Pre-specified sub-group analyses were conducted according to participants’ sex (males vs. females), baseline deseasonalized 25(OH)D concentration (<10 vs. ≥10 ng/mL) and calcium intake (<500 vs. ≥500 mg/day). The primary P-values for sub-group analyses were the overall P values, i.e. those associated with the interaction between follow-up timepoint and treatment allocation.

### Role of the funding source

The funder of the study had no role in study design, data collection, data analysis, data interpretation or writing of this report.

## RESULTS

### PARTICIPANTS

11475 children were invited to participate in the study from September 2015 through March 2017, of whom 9814 underwent QFT testing: 8851 QFT-negative children were randomly assigned to receive vitamin D_3_ or placebo (Fig. 1). 726/4418 participants assigned to the vitamin D arm and 739/4433 participants assigned to the placebo arm participated in the BMD sub-study; 50 participants from each arm contributed serum samples to the bone biochemistry sub-study (Fig. 1). Table 1 presents baseline characteristics of children in the main trial, the BMD sub-study and the bone biochemistry sub-study, overall and by study arm. Mean age was 9.4 years and 49.3% were female. Mean baseline serum 25(OH)D concentration was 11.9 ng/mL; 95.6% of participants had 25(OH)D levels <20 ng/mL and 31.8% had 25(OH)D levels <10 ng/mL. Baseline characteristics were balanced between those randomized to vitamin D vs. placebo for participants in the main trial, the BMD sub-study and the bone biochemistry sub-study. Mean end-trial serum 25(OH)D was 31.0 ng/ml in the treatment group and 10.7 in the placebo group (mean difference: 20.3 ng/mL, 95%CI 19.9-20.6). End-study 25(OH)D levels were ≥20 ng/mL in 89.8% of participants randomized to vitamin D vs. 5.6% of those randomized to placebo.

**Figure 1:**
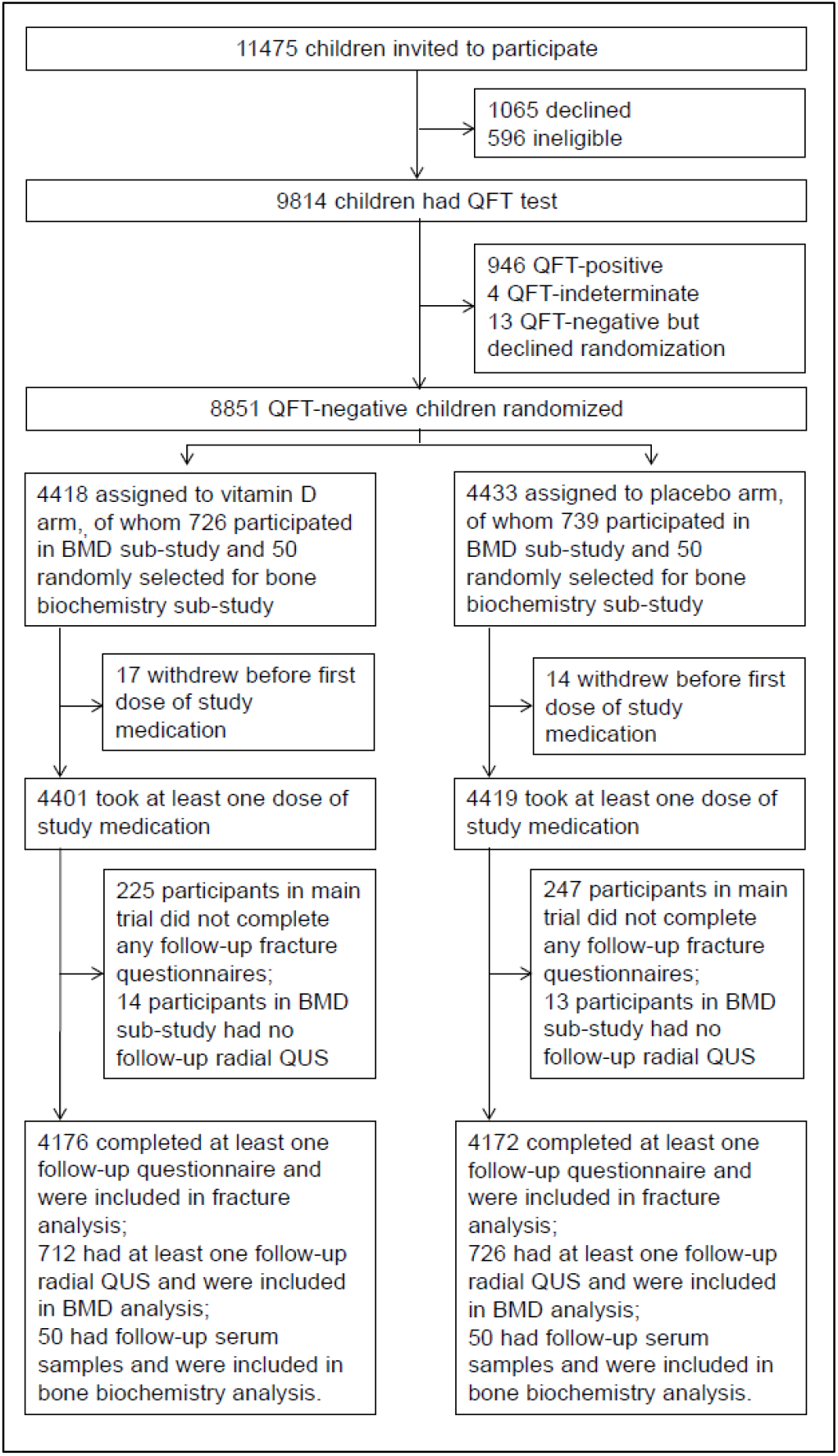
Participant flow

**Table 1:** Participants’ baseline characteristics by allocation: main trial, BMD sub-study and bone biochemistry sub-study

|  |  | Main trial |  |  | BMD sub-study |  |  | Bone biochemistry sub-study |  |  |
| --- | --- | --- | --- | --- | --- | --- | --- | --- | --- | --- |
|  |  | (n=8,851) |  |  | (n=1,465 subset) |  |  | (n=100 subset) |  |  |
|  |  | Overall<br>(n=8,851) | Vitamin D<br>(n=4,418) | Placebo<br>(n=4,433) | Overall<br>(n=1,465) | Vitamin D<br>(n=726) | Placebo<br>(n=739) | Overall<br>(n=100) | Vitamin D<br>(n=50) | Placebo<br>(n=50) |
| Mean age, years (s.d.) |  | 9.4 (1.6) | 9.4 (1.6) | 9.4 (1.6) | 9.5 (1.6) | 9.5 (1.6) | 9.5 (1.5) | 9.2 (1.5) | 9.4 (1.5) | 9 (1.5) |
| Female sex, n (%) |  | 4125 (49.4) | 2025 (48.5) | 2100 (50.3) | 717 (48.9) | 361 (49.7) | 356 (48.2) | 51 (51.0) | 27 (54.0) | 24 (48.0) |
| Ethnic origin | Khal'kh, n (%) | 7701 (92.2) | 3838 (91.9) | 3863 (92.6) | 1346 (91.9) | 660 (90.9) | 686 (92.8) | 87 (87.0) | 43 (86.0) | 44 (88.0) |
|  | Other, n (%) | 647 (7.8) | 338 (8.1) | 309 (7.4) | 119 (8.1) | 66 (9.1) | 53 (7.2) | 13 (13.0) | 7 (14.0) | 6 (12.0) |
| Parental education<sup>(1)</sup> | Secondary school or lower, n (%) | 4579 (54.9) | 2267 (54.3) | 2312 (55.4) | 882 (60.2) | 432 (59.5) | 450 (60.9) | 61 (61.0) | 32 (64.0) | 29 (58.0) |
|  | University/polytechnic, n(%) | 3769 (45.1) | 1909 (45.7) | 1860 (44.6) | 583 (39.8) | 294 (40.5) | 289 (39.1) | 39 (39.0) | 18 (36.0) | 21 (42.0) |
| Type of residence | Ger (Yurt) , n (%) | 3091 (37) | 1555 (37.2) | 1536 (36.8) | 613 (41.8) | 319 (43.9) | 294 (39.8) | 47 (47.0) | 25 (50.0) | 22 (44.0) |
|  | House without central heating, n (%) | 3231 (38.7) | 1586 (38) | 1645 (39.4) | 624 (42.6) | 296 (40.8) | 328 (44.4) | 38 (38.0) | 16 (32.0) | 22 (44.0) |
|  | House with central heating, n (%) | 2026 (24.3) | 1035 (24.8) | 991 (23.8) | 228 (15.6) | 111 (15.3) | 117 (15.8) | 15 (15.0) | 9 (18.0) | 6 (12.0) |
| Home ownership, n (%) |  | 6594 (79) | 3310 (79.3) | 3284 (78.7) | 1111 (75.8) | 552 (76) | 559 (75.6) | 81 (81.0) | 43 (86.0) | 38 (76.0) |
| Mean monthly household income, US dollars (s.d.)<sup>(2)</sup> |  | 842.8 (537.3) | 847.4 (549.5) | 838.1 (524.7) | 766.8 (437.4) | 776.8 (433.7) | 757.1 (441.1) | 745.5 (510.7) | 746.6 (528) | 744.4 (498.2) |
| Calcium intake<sup>(2)</sup> | <500 mg/day, n (%) | 5191 (62.2) | 2582 (61.8) | 2609 (62.5) | 888 (60.6) | 451 (62.1) | 437 (59.1) | 62 (62.0) | 30 (60.0) | 32 (64.0) |
|  | ≥500 mg/day, n (%) | 2899 (34.7) | 1471 (35.2) | 1428 (34.2) | 486 (33.2) | 236 (32.5) | 250 (33.8) | 36 (36.0) | 19 (38.0) | 17 (34.0) |
| Mean BMI-for-age z-score (s.d.)<sup>(2)</sup> |  | 0.2 (1.1) | 0.2 (1) | 0.2 (1.1) | 0.1 (1) | 0.1 (1.1) | 0.1 (1) | 0.2 (0.9) | 0.1 (0.9) | 0.3 (1) |
| Mean height-for-age z-score (s.d.)<sup>(2)</sup> |  | -0.3 (1) | -0.3 (1) | -0.3 (1) | -0.3 (1) | -0.3 (1) | -0.4 (0.9) | -0.4 (1.1) | -0.4 (1.1) | -0.5 (1.1) |
| Mean serum 25(OH)D, ng/ml (s.d.)<sup>(2,4)</sup> |  | 11.9 (4.2) | 11.8 (4.2) | 11.9 (4.2) | 11.6 (4) | 11.6 (4) | 11.7 (3.9) | 11.9 (4.3) | 11.8 (4.5) | 11.9 (4.1) |
| Serum 25(OH)D<sup>(2,4)</sup> | <10 ng/ml, n (%) | 2664 (31.9) | 1323 (31.7) | 1341 (32.2) | 526 (35.9) | 254 (35) | 272 (36.8) | 34 (34.0) | 15 (30.0) | 19 (38.0) |
|  | 10-19.9 ng/ml, n (%) | 5311 (63.7) | 2671 (64) | 2640 (63.3) | 882 (60.2) | 442 (60.9) | 440 (59.5) | 60 (60.0) | 31 (62.0) | 29 (58.0) |
|  | 20-29.9 ng/ml, n (%) | 381 (4.3) | 174 (4.2) | 185 (4.4) | 56 (3.8) | 29 (4) | 27 (3.7) | 6 (6.0) | 4 (8.0) | 2 (4.0) |
|  | ≥30 ng/ml, n (%) | 10 (0.1) | 7 (0.2) | 3 (0.1) | 1 (0.1) | 1 (0.1) | (0) | 0 (0.0) | 0 (0.0) | 0 (0.0) |
| Mean radial QUS z-score (s.d.) | -- | -- | -- | -- | -0.6 (1.1) | -0.5 (1.1) | -0.6 (1.1) | -- | -- | -- |
| Mean radial speed of sound, m/s (s.d.) | -- | -- | -- |  | 3635.1 (121.8) | 3638.4 (122.8) | 3631.9 (120.8) | 3619.3 (125.1) | 3609.5 (133.2) | 3629 (116.9) |
| Mean adjusted serum calcium concentration, mmol/L (s.d.) | -- | -- | -- | -- | -- | -- | -- | 2.23 (0.10) | 2.22 (0.12) | 2.24 (0.08) |
| Mean serum PTH concentration, pmol/L (s.d.) | -- | -- | -- | -- | -- | -- | -- | 52.1 (35.8) | 53.5 (46.1) | 50.7 (21.6) |
| Mean serum total ALP concentration, IU/L (s.d.) | -- | -- | -- | -- | -- | -- | -- | 291.8 (85.7) | 285.7 (86.4) | 298 (85.4) |
| Mean serum bone-specific ALP concentration, IU/L (s.d.) | -- | -- | -- | -- | -- | -- | -- | 26.4 (8.8) | 26.6 (9.1) | 26.2 (8.6) |

**Abbreviations:** 25(OH)D, 25-hydroxyvitamin D. ALP, alkaline phosphatase. BMI, body mass index. S.d., standard deviation. QUS, quantitative ultrasound. US, United States.
**Footnotes.** (1) Highest educational level attained by either parent; (2) Missing values: for fracture study, household income missing for 1 participant in each arm; calcium intake missing for 365 participants in vitamin D arm and 376 participants in placebo arm; baseline BMI-for-age z-score missing for 1 participant in placebo arm; baseline height-for-age z-score missing for 1 participant in placebo arm; baseline serum 25(OH)D missing for 1 participant in vitamin D arm and 4 participants in placebo arm; for BMD sub-study, calcium intake missing for 39 participants in vitamin D arm and 52 participants in placebo arm. For bone biochemistry sub-study, calcium intake missing for 1 participant in vitamin D arm and 1 participant in placebo arm. (3) Defined as one or more people in the household smoking indoors; (4) baseline 25(OH)D values deseasonalized.

### OUTCOMES

A total of 677 fractures arose in 521 participants during 25407.7 person-years of follow-up, of which 461 affected the upper limb, 163 affected the lower limb and 53 occurred at other sites (Table S1, Supplementary Material). The proportions of children reporting one or more fractures were similar for those randomized to vitamin D vs. placebo (268/4176 [6.4%] vs. 253/4172 [6.1%], respectively; adjusted risk ratio 1.10, 95% CI 0.93 to 1.29, P=0.27; Table 2). Sub-group analyses for this outcome showed no evidence of effect modification by sex, baseline 25(OH)D concentration or calcium intake (P values for interaction all >0.05; Table 2). No inter-arm differences in proportions of participants reporting one or more fractures were seen for X-ray confirmed or plaster cast-treated events; for fractures arising at different sites; or for fractures associated with different degrees of trauma (Table S2, Supplementary Material). Among participants in the BMD sub-study, allocation to vitamin D vs. placebo did not influence radial BMD z-score or speed-of-sound, either overall or in sub-groups defined by sex, baseline 25(OH)D concentration or calcium intake (Table 3, Table S3 Supplementary Material). Among participants contributing serum to the bone biochemistry sub-study, participants randomized to vitamin D vs. placebo had higher 25(OH)D concentrations (mean difference 20.1 ng/mL, 95% CI 19.8 to 20.4 ng/mL) and lower PTH concentrations (mean difference −13.6 pmol/L, 95% CI −23.5 to −3.7 pmol/L). The effect of vitamin D on PTH concentrations was modified both by baseline 25(OH)D concentration and by calcium intake, with greater vitamin D-induced reductions in PTH seen in participants having baseline 25(OH)D concentrations <10 vs. ≥10 ng/mL (P for interaction 0.04) and calcium intakes <500 vs. ≥500 mg/day (P for interaction 0.03). Allocation to vitamin D vs. placebo did not influence adjusted serum calcium concentrations overall, but the effect of allocation was modified by baseline 25(OH)D concentration, with a greater vitamin D-induced increase seen in participants having baseline 25(OH)D concentrations <10 vs. ≥10 ng/mL (P for interaction 0.01). No overall effect of allocation was seen on total serum ALP or BALP concentrations, but the effect of vitamin D on both parameters was modified by baseline 25(OH)D concentrations, with greater vitamin D-induced reductions seen in participants having baseline 25(OH)D concentrations <10 vs. ≥10 ng/mL (P for interaction ≤0.04). The effect of vitamin D on BALP was also modified by calcium intake, with greater vitamin D-induced reductions seen in those with calcium intake <500 vs ≥500 mg/day (P for interaction =0.02).

**Table 2.** Proportion of participants reporting one or more bone fractures of any type by allocation, overall and by sub-group

|  |  | Vitamin D arm<br>(n=4176) | Placebo arm<br>(n=4172) | Adjusted risk ratio<br>(95% CI) <sup>(1)</sup> | P | P for<br>interaction <sup>(2)</sup> |
| --- | --- | --- | --- | --- | --- | --- |
| Overall |  | 268 (6.4%) | 253 (6.1%) | 1.10 (0.93 to 1.29) | 0.27 |  |
| By sex | Male | 183 (8.5%) | 164 (7.9%) | 1.11 (0.91 to 1.35) | 0.30 | 0.60 |
|  | Female | 85 (4.2%) | 89 (4.2%) | 0.99 (0.75 to 1.3) | 0.92 |  |
| By baseline 25(OH)D<br>concentration <sup>(3)</sup> | <10 ng/mL | 82 (6.2%) | 73 (5.4%) | 1.13 (0.84 to 1.5) | 0.42 | 0.67 |
|  | ≥10 ng/mL | 186 (6.5%) | 180 (6.4%) | 1.08 (0.90 to 1.31) | 0.41 |  |
| By calcium intake | <500 mg/day | 169 (6.5%) | 162 (6.2%) | 1.11 (0.91 to 1.36) | 0.29 | 0.94 |
|  | ≥500 mg/day | 91 (6.2%) | 84 (5.9%) | 1.07 (0.81 to 1.41) | 0.63 |  |

**Abbreviations:** 25(OH)D<sub>3</sub>, 25-hydroxyvitamin D<sub>3</sub>. CI, confidence interval.
**Footnotes.** 1, adjusted for random effects of school and individual. 2, P-value for treatment-by-subgroup interaction. 3, deseasonalized values.

**Table 3.**
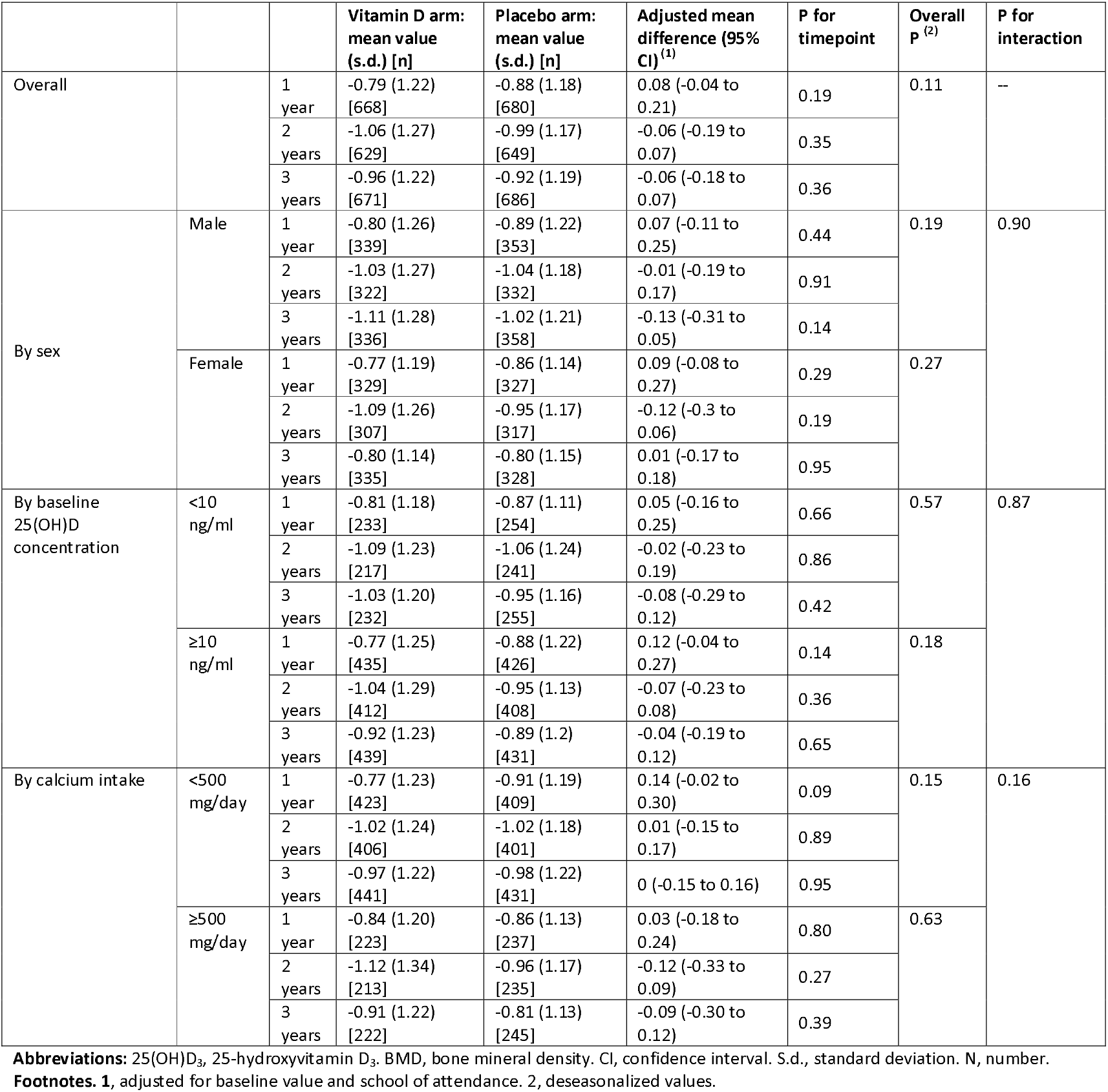
Mean radial BMD z-scores at 1-, 2- and 3-year follow-up by allocation: overall and by sub-group

|  |  |  | Vitamin D arm:<br>mean value<br>(s.d.) [n] | Placebo arm:<br>mean value<br>(s.d.) [n] | Adjusted mean<br>difference (95%<br>CI)<sup>(1)</sup> | P for<br>timepoint | Overall<br>p<sup>(2)</sup> | P for<br>interaction |
| --- | --- | --- | --- | --- | --- | --- | --- | --- |
| Overall |  | 1<br>year | -0.79 (1.22)<br>[668] | -0.88 (1.18)<br>[680] | 0.08 (-0.04 to<br>0.21) | 0.19 | 0.11 | -- |
|  |  | 2<br>years | -1.06 (1.27)<br>[629] | -0.99 (1.17)<br>[649] | -0.06 (-0.19 to<br>0.07) | 0.35 |  |  |
|  |  | 3<br>years | -0.96 (1.22)<br>[671] | -0.92 (1.19)<br>[686] | -0.06 (-0.18 to<br>0.07) | 0.36 |  |  |
| By sex | Male | 1<br>year | -0.80 (1.26)<br>[339] | -0.89 (1.22)<br>[353] | 0.07 (-0.11 to<br>0.25) | 0.44 | 0.19 | 0.90 |
|  |  | 2<br>years | -1.03 (1.27)<br>[322] | -1.04 (1.18)<br>[332] | -0.01 (-0.19 to<br>0.17) | 0.91 |  |  |
|  |  | 3<br>years | -1.11 (1.28)<br>[336] | -1.02 (1.21)<br>[358] | -0.13 (-0.31 to<br>0.05) | 0.14 |  |  |
|  | Female | 1<br>year | -0.77 (1.19)<br>[329] | -0.86 (1.14)<br>[327] | 0.09 (-0.08 to<br>0.27) | 0.29 | 0.27 |  |
|  |  | 2<br>years | -1.09 (1.26)<br>[307] | -0.95 (1.17)<br>[317] | -0.12 (-0.3 to<br>0.06) | 0.19 |  |  |
|  |  | 3<br>years | -0.80 (1.14)<br>[335] | -0.80 (1.15)<br>[328] | 0.01 (-0.17 to<br>0.18) | 0.95 |  |  |
| By baseline<br>25(OH)D<br>concentration | <10<br>ng/ml | 1<br>year | -0.81 (1.18)<br>[233] | -0.87 (1.11)<br>[254] | 0.05 (-0.16 to<br>0.25) | 0.66 | 0.57 | 0.87 |
|  |  | 2<br>years | -1.09 (1.23)<br>[217] | -1.06 (1.24)<br>[241] | -0.02 (-0.23 to<br>0.19) | 0.86 |  |  |
|  |  | 3<br>years | -1.03 (1.20)<br>[232] | -0.95 (1.16)<br>[255] | -0.08 (-0.29 to<br>0.12) | 0.42 |  |  |
|  | ≥10<br>ng/ml | 1<br>year | -0.77 (1.25)<br>[435] | -0.88 (1.22)<br>[426] | 0.12 (-0.04 to<br>0.27) | 0.14 | 0.18 |  |
|  |  | 2<br>years | -1.04 (1.29)<br>[412] | -0.95 (1.13)<br>[408] | -0.07 (-0.23 to<br>0.08) | 0.36 |  |  |
|  |  | 3<br>years | -0.92 (1.23)<br>[439] | -0.89 (1.2)<br>[431] | -0.04 (-0.19 to<br>0.12) | 0.65 |  |  |
| By calcium intake | <500<br>mg/day | 1<br>year | -0.77 (1.23)<br>[423] | -0.91 (1.19)<br>[409] | 0.14 (-0.02 to<br>0.30) | 0.09 | 0.15 | 0.16 |
|  |  | 2<br>years | -1.02 (1.24)<br>[406] | -1.02 (1.18)<br>[401] | 0.01 (-0.15 to<br>0.17) | 0.89 |  |  |
|  |  | 3<br>years | -0.97 (1.22)<br>[441] | -0.98 (1.22)<br>[431] | 0 (-0.15 to 0.16) | 0.95 |  |  |
|  | ≥500<br>mg/day | 1<br>year | -0.84 (1.20)<br>[223] | -0.86 (1.13)<br>[237] | 0.03 (-0.18 to<br>0.24) | 0.80 | 0.63 |  |
|  |  | 2<br>years | -1.12 (1.34)<br>[213] | -0.96 (1.17)<br>[235] | -0.12 (-0.33 to<br>0.09) | 0.27 |  |  |
|  |  | 3<br>years | -0.91 (1.22)<br>[222] | -0.81 (1.13)<br>[245] | -0.09 (-0.30 to<br>0.12) | 0.39 |  |  |

**Abbreviations:** 25(OH)D<sub>3</sub>, 25-hydroxyvitamin D<sub>3</sub>. BMD, bone mineral density. CI, confidence interval. S.d., standard deviation. N, number.
**Footnotes.** 1, adjusted for baseline value and school of attendance. 2, deseasonalized values.

## DISCUSSION

We present findings of the largest RCT to investigate effects of vitamin D supplementation on risk of bone fracture in children. Profound vitamin D deficiency and low calcium intakes were common among participants at baseline, and weekly oral administration of 14,000 IU vitamin D_3_ was effective in elevating serum 25(OH)D concentrations into the high physiologic range. In a subset of participants contributing serum for biochemical analyses, this was associated with reduced serum PTH concentrations (irrespective of baseline vitamin D status) and suppression of serum concentrations of total ALP and BALP in participants with baseline serum 25(OH)D concentration <10 ng/mL. However, the intervention did not influence fracture risk or radial bone mineral density, either in the study population as a whole or in sub-groups of participants with low baseline vitamin D status or low calcium intake.

Null results in our trial for outcomes of fracture incidence and BMD are consistent with those of our sister trial conducted in Cape Town schoolchildren^14^ and with those of RCTs conducted in adults, which tend to show that vitamin D is only effective in reducing fracture risk when administered together with calcium supplementation.^15^ The lack of concomitant calcium supplementation in our trial may have been particularly impactful on the outcome, given that 62.2% of participants had calcium intakes less 500 mg/day-the minimum recommended during childhood and adolescence.^16^ However, no effect of vitamin D on fracture risk or BMD was seen even in the sub-group of participants with higher calcium intake. Suppression of serum PTH concentrations in response to vitamin D supplementation likely reflects correction of secondary hyperparathyroidism associated with participants’ very low vitamin D status at baseline. Vitamin D-induced suppression of BALP in participants with the lowest baseline 25(OH)D concentrations might have been expected to be associated with increased bone formation in this sub-group: however, we saw no inter-arm differences in BMD or fracture risk even when analysis was restricted to those with baseline 25(OH)D concentrations <10 ng/mL.

Our study has several strengths. The double-blind RCT design is the gold standard for evaluating effectiveness of an intervention, as it minimizes the potential for bias and confounding to operate. Vitamin D deficiency was prevalent at baseline, and the intervention was effective in elevating serum 25(OH)D concentrations into the high physiologic range. The 3-year duration of the study provided ample time for any effects of vitamin D to manifest. Fracture incidence was high at 266 events per 10,000 children per year – more than double that reported in a Norwegian cohort.^3^ Together with the large sample size, this provided a high degree of statistical power to detect an effect of the intervention. The lower bound of the 95% confidence interval for the adjusted risk ratio was 0.93, indicating 97.5% certainty that any reduction in risk of experiencing a fracture is no greater than 7%: our null findings can therefore be regarded as definitive. The consistent lack of signal for radial BMD and fracture outcomes enhances confidence that our null results are likely to be valid.

Our study also has some limitations. We employed radial QUS to measure BMD rather than DXA, which is the gold standard methodology. However, the low cost, simplicity of performance, mobility and lack of exposure to ionizing radiation made this alternative attractive. We highlight that QUS-determined BMD associates with fracture risk in adults,^17^ and that low SOS values associate with increased risk of fracture in children.^18^ We only investigated a single dose of vitamin D, which was given at weekly intervals without concomitant calcium supplementation: thus, our findings do not exclude the possibility that another dosing regimen, with or without concomitant calcium supplementation, may be effective.

In conclusion, this phase 3 multicenter trial, conducted in schoolchildren with low baseline vitamin D status and low calcium intake, reports that weekly oral administration of 14,000 IU vitamin D3 for 3 years was effective in elevating serum 25(OH)D concentrations into the physiologic range. This resulted in suppression of PTH concentrations overall, and reduction of total and bone-specific ALP concentrations in participants with low baseline vitamin D status. However, these biochemical changes were not associated with reduced fracture risk or increased bone mineral density.

## Data Availability

Anonymised data may be requested from the corresponding authors to be shared subject to terms of IRB approval.

## CONTRIBUTORS

DG and ARM conceived the study and contributed to study design and protocol development. DG led on trial implementation, with support from UB, ET, SE, C-E A, NY, BD, MA, ET, BO, BJ and ED. BD and MA performed and supervised the conduct of biochemical assays. DG, PK and ARM drafted the statistical analysis plan. DG, UB, ET, SE, BD and MA managed data. PK conducted statistical analyses. ARM wrote the first draft of the trial report. All authors made substantive comments thereon and approved the final version for submission.

## DECLARATION OF INTERESTS

ARM declares receipt of funding in the last 36 months to support vitamin D research from the following companies who manufacture or sell vitamin D supplements: Pharma Nord Ltd, DSM Nutritional Products Ltd, Thornton & Ross Ltd and Hyphens Pharma Ltd. ARM also declares receipt of vitamin D capsules for clinical trial use from Pharma Nord Ltd, Synergy Biologics Ltd and Cytoplan Ltd; support for attending meetings from Pharma Nord Ltd and Abiogen Pharma Ltd; receipt of consultancy fees from DSM Nutritional Products Ltd and Qiagen Ltd; receipt of a speaker fee from the Linus Pauling Institute; participation on Data and Safety Monitoring Boards for the VITALITY trial (Vitamin D for Adolescents with HIV to reduce musculoskeletal morbidity and immunopathology, Pan African Clinical Trials Registry ref PACTR20200989766029) and the Trial of Vitamin D and Zinc Supplementation for Improving Treatment Outcomes Among COVID-19 Patients in India (http://ClinicalTrials.gov ref NCT04641195); and unpaid work as a Programme Committee member for the Vitamin D Workshop. All other authors declare that they have no competing interests.

## ACKNOWLEDGEMENTS

This study was supported by an award from the United States National Institutes of Health, ref. 1R01HL122624-01. We thank all the children who participated in the trial, and their parents and guardians. We also thank independent members of the Data Safety Monitoring Board (Prof S.M. Fortune and Dr P.L. Williams, Harvard T.H. Chan School of Public Health; Profs M.F. Holick and C.R. Horsburgh, Boston University; Prof P. Enkhbaatar, University of Texas; and Dr. E. Chadraa, Minnesota State University); independent members of the Trial Steering Committee (Profs W.C. Willett, Edward L. Giovannucci and B.R. Bloom, Harvard T.H. Chan School of Public Health; Dr N. Naranbat, Gyals Medical Laboratory, Ulaanbaatar; and late Dr D. Malchinkhuu, National Center for Maternal and Child Health of Mongolia); board members at the Mongolian Health Initiative (Dr J. Tuyatsetseg, Mongolian University of Science and Technology; Dr J. Amarsanaa, Happy Veritas Laboratory, Ulaanbaatar; Drs P. Erkhembulgan and G. Batbaatar, Mongolian National University of Medical Sciences; Prof M.C. Elliott, Harvard University; and Mr. Ts. Munh-Orgil, Member of the Mongolian Parliament); and the following individuals for advice and helpful discussions: Dr Winthrop Burr (Anadyne Psychotherapy Inc) and and Dr. Masae Kawamura at Qiagen USA.

---

**Table 4.**
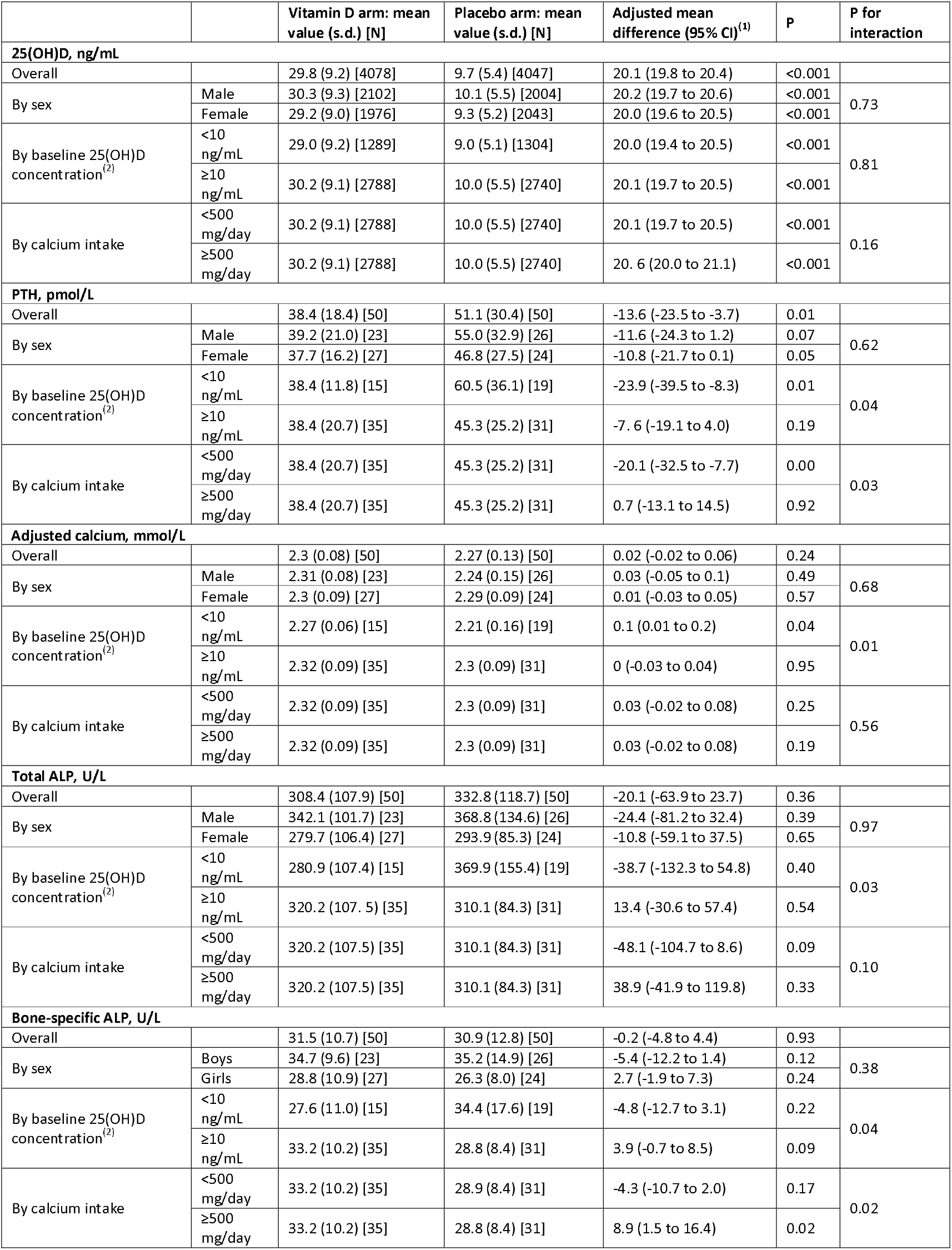
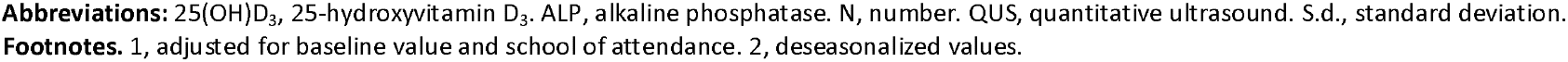
Serum concentrations of 25(OH)D, adjusted calcium, PTH, total ALP and bone-specific ALP at 3-year follow-up by allocation: overall and by sub-group

|  |  | Vitamin D arm: mean value (s.d.) [N] | Placebo arm: mean value (s.d.) [N] | Adjusted mean difference (95% CI)<sup>(1)</sup> | P | P for interaction |
| --- | --- | --- | --- | --- | --- | --- |
| 25(OH)D, ng/mL |  |  |  |  |  |  |
| Overall |  | 29.8 (9.2) [4078] | 9.7 (5.4) [4047] | 20.1 (19.8 to 20.4) | <0.001 |  |
| By sex | Male | 30.3 (9.3) [2102] | 10.1 (5.5) [2004] | 20.2 (19.7 to 20.6) | <0.001 | 0.73 |
|  | Female | 29.2 (9.0) [1976] | 9.3 (5.2) [2043] | 20.0 (19.6 to 20.5) | <0.001 |  |
| By baseline 25(OH)D concentration<sup>(2)</sup> | <10 ng/mL | 29.0 (9.2) [1289] | 9.0 (5.1) [1304] | 20.0 (19.4 to 20.5) | <0.001 | 0.81 |
|  | ≥10 ng/mL | 30.2 (9.1) [2788] | 10.0 (5.5) [2740] | 20.1 (19.7 to 20.5) | <0.001 |  |
| By calcium intake | <500 mg/day | 30.2 (9.1) [2788] | 10.0 (5.5) [2740] | 20.1 (19.7 to 20.5) | <0.001 | 0.16 |
|  | ≥500 mg/day | 30.2 (9.1) [2788] | 10.0 (5.5) [2740] | 20.6 (20.0 to 21.1) | <0.001 |  |
| PTH, pmol/L |  |  |  |  |  |  |
| Overall |  | 38.4 (18.4) [50] | 51.1 (30.4) [50] | -13.6 (-23.5 to -3.7) | 0.01 |  |
| By sex | Male | 39.2 (21.0) [23] | 55.0 (32.9) [26] | -11.6 (-24.3 to 1.2) | 0.07 | 0.62 |
|  | Female | 37.7 (16.2) [27] | 46.8 (27.5) [24] | -10.8 (-21.7 to 0.1) | 0.05 |  |
| By baseline 25(OH)D concentration<sup>(2)</sup> | <10 ng/mL | 38.4 (11.8) [15] | 60.5 (36.1) [19] | -23.9 (-39.5 to -8.3) | 0.01 | 0.04 |
|  | ≥10 ng/mL | 38.4 (20.7) [35] | 45.3 (25.2) [31] | -7.6 (-19.1 to 4.0) | 0.19 |  |
| By calcium intake | <500 mg/day | 38.4 (20.7) [35] | 45.3 (25.2) [31] | -20.1 (-32.5 to -7.7) | 0.00 | 0.03 |
|  | ≥500 mg/day | 38.4 (20.7) [35] | 45.3 (25.2) [31] | 0.7 (-13.1 to 14.5) | 0.92 |  |
| Adjusted calcium, mmol/L |  |  |  |  |  |  |
| Overall |  | 2.3 (0.08) [50] | 2.27 (0.13) [50] | 0.02 (-0.02 to 0.06) | 0.24 |  |
| By sex | Male | 2.31 (0.08) [23] | 2.24 (0.15) [26] | 0.03 (-0.05 to 0.1) | 0.49 | 0.68 |
|  | Female | 2.3 (0.09) [27] | 2.29 (0.09) [24] | 0.01 (-0.03 to 0.05) | 0.57 |  |
| By baseline 25(OH)D concentration<sup>(2)</sup> | <10 ng/mL | 2.27 (0.06) [15] | 2.21 (0.16) [19] | 0.1 (0.01 to 0.2) | 0.04 | 0.01 |
|  | ≥10 ng/mL | 2.32 (0.09) [35] | 2.3 (0.09) [31] | 0 (-0.03 to 0.04) | 0.95 |  |
| By calcium intake | <500 mg/day | 2.32 (0.09) [35] | 2.3 (0.09) [31] | 0.03 (-0.02 to 0.08) | 0.25 | 0.56 |
|  | ≥500 mg/day | 2.32 (0.09) [35] | 2.3 (0.09) [31] | 0.03 (-0.02 to 0.08) | 0.19 |  |
| Total ALP, U/L |  |  |  |  |  |  |
| Overall |  | 308.4 (107.9) [50] | 332.8 (118.7) [50] | -20.1 (-63.9 to 23.7) | 0.36 |  |
| By sex | Male | 342.1 (101.7) [23] | 368.8 (134.6) [26] | -24.4 (-81.2 to 32.4) | 0.39 | 0.97 |
|  | Female | 279.7 (106.4) [27] | 293.9 (85.3) [24] | -10.8 (-59.1 to 37.5) | 0.65 |  |
| By baseline 25(OH)D concentration<sup>(2)</sup> | <10 ng/mL | 280.9 (107.4) [15] | 369.9 (155.4) [19] | -38.7 (-132.3 to 54.8) | 0.40 | 0.03 |
|  | ≥10 ng/mL | 320.2 (107.5) [35] | 310.1 (84.3) [31] | 13.4 (-30.6 to 57.4) | 0.54 |  |
| By calcium intake | <500 mg/day | 320.2 (107.5) [35] | 310.1 (84.3) [31] | -48.1 (-104.7 to 8.6) | 0.09 | 0.10 |
|  | ≥500 mg/day | 320.2 (107.5) [35] | 310.1 (84.3) [31] | 38.9 (-41.9 to 119.8) | 0.33 |  |
| Bone-specific ALP, U/L |  |  |  |  |  |  |
| Overall |  | 31.5 (10.7) [50] | 30.9 (12.8) [50] | -0.2 (-4.8 to 4.4) | 0.93 |  |
| By sex | Boys | 34.7 (9.6) [23] | 35.2 (14.9) [26] | -5.4 (-12.2 to 1.4) | 0.12 | 0.38 |
|  | Girls | 28.8 (10.9) [27] | 26.3 (8.0) [24] | 2.7 (-1.9 to 7.3) | 0.24 |  |
| By baseline 25(OH)D concentration<sup>(2)</sup> | <10 ng/mL | 27.6 (11.0) [15] | 34.4 (17.6) [19] | -4.8 (-12.7 to 3.1) | 0.22 | 0.04 |
|  | ≥10 ng/mL | 33.2 (10.2) [35] | 28.8 (8.4) [31] | 3.9 (-0.7 to 8.5) | 0.09 |  |
| By calcium intake | <500 mg/day | 33.2 (10.2) [35] | 28.9 (8.4) [31] | -4.3 (-10.7 to 2.0) | 0.17 | 0.02 |
|  | ≥500 mg/day | 33.2 (10.2) [35] | 28.8 (8.4) [31] | 8.9 (1.5 to 16.4) | 0.02 |  |

**Abbreviations:** 25(OH)D<sub>3</sub>, 25-hydroxyvitamin D<sub>3</sub>. ALP, alkaline phosphatase. N, number. QUS, quantitative ultrasound. S.d., standard deviation. **Footnotes.** 1, adjusted for baseline value and school of attendance. 2, deseasonalized values.

## Notes

### Clinical Trial

NCT04641195

### Author Declarations

The study was approved by institutional review boards of the Mongolian Ministry of Health, Mongolian National University, and Harvard T. H. Chan School of Public Health (IRB # 14-0513).

## References

1. Cooper C, Dennison EM, Leufkens HG, Bishop N, van Staa TP. Epidemiology of childhood fractures in Britain: a study using the general practice research database. J Bone Miner Res 2004; 19(12): 1976–81.

2. Hedstrom EM, Svensson O, Bergstrom U, Michno P. Epidemiology of fractures in children and adolescents. Acta Orthop 2010; 81(1): 148–53.

3. Kopjar B, Wickizer TM. Fractures among children: incidence and impact on daily activities. Inj Prev 1998; 4(3): 194–7.

4. Mace SE, Gerardi MJ, Dietrich AM, et al. Injury prevention and control in children. Annals of emergency medicine 2001; 38(4): 405–14.

5. Proia P, Amato A, Drid P, Korovljev D, Vasto S, Baldassano S. The Impact of Diet and Physical Activity on Bone Health in Children and Adolescents. Front Endocrinol (Lausanne) 2021; 12: 704647.

6. Holick MF. Vitamin D deficiency. N Engl J Med 2007; 357(3): 266–81.

7. Moon RJ, Harvey NC, Davies JH, Cooper C. Vitamin D and skeletal health in infancy and childhood. Osteoporos Int 2014; 25(12): 2673–84.

8. GBD Fracture Collaborators. Global, regional, and national burden of bone fractures in 204 countries and territories, 1990-2019: a systematic analysis from the Global Burden of Disease Study 2019. Lancet Healthy Longev 2021; 2(9): e580–e92.

9. Bater J, Bromage S, Jambal T, et al. Prevalence and Determinants of Vitamin D Deficiency in 9595 Mongolian Schoolchildren: A Cross-Sectional Study. Nutrients 2021; 13(11).

10. Ganmaa D, Uyanga B, Zhou X, et al. Vitamin D Supplements for Prevention of Tuberculosis Infection and Disease. N Engl J Med 2020; 383(4): 359–68.

11. Ganmaa D, Bromage S, Khudyakov P, Erdenenbaatar S, Delgererekh B, Martineau AR. Influence of Vitamin D Supplementation on Growth, Body Composition, and Pubertal Development Among School-aged Children in an Area With a High Prevalence of Vitamin D Deficiency: A Randomized Clinical Trial. JAMA pediatrics 2023; 177(1): 32–41.

12. Bromage S, Daria T, Lander RL, et al. Diet and Nutrition Status of Mongolian Adults. Nutrients 2020; 12(5).

13. BeamMed Ltd. User guide: Sunlight MiniOmni™ Ultrasound Bone Sonometer. Document Number DUM-0090, Revision 01. 2014. https://www.beammed.com/wp-content/uploads/2022/02/DUM-0081-MiniOmni-UG-rev.-08-FNL-1.pdf (accessed 05/12/2023.

14. Middelkoop K, Micklesfield LK, Walker N, et al. Influence of vitamin D supplementation on bone mineral content, bone turnover markers and fracture risk in South African schoolchildren: multicentre double-blind randomised placebo-controlled trial (ViDiKids). MedRxiv 2023.

15. Chakhtoura M, Bacha DS, Gharios C, et al. Vitamin D Supplementation and Fractures in Adults: A Systematic Umbrella Review of Meta-Analyses of Controlled Trials. J Clin Endocrinol Metab 2022; 107(3): 882–98.

16. Munns CF, Shaw N, Kiely M, et al. Global Consensus Recommendations on Prevention and Management of Nutritional Rickets. J Clin Endocrinol Metab 2016; 101(2): 394–415.

17. Moayyeri A, Adams JE, Adler RA, et al. Quantitative ultrasound of the heel and fracture risk assessment: an updated meta-analysis. Osteoporos Int 2012; 23(1): 143–53.

18. Schalamon J, Singer G, Schwantzer G, Nietosvaara Y. Quantitative ultrasound assessment in children with fractures. J Bone Miner Res 2004; 19(8): 1276–9.

