## Supplementary Appendix for "Influence of vitamin D supplementation on fracture risk, bone mineral density and bone biochemistry in Mongolian schoolchildren: multicenter double-blind randomized placebo-controlled trial"

**Supplementary Material**

### **Randomization and Masking**

Eligible participants were individually randomized to receive a weekly capsule containing vitamin D_3_ or placebo for three years, with a one-to-one allocation ratio. Randomization was stratified by school of attendance (a potential predictor of risk of QFT conversion) as follows. Prior to the start of recruitment, Dr Polyna Khudyakov (Harvard School of Public Health) prepared a single school randomization list comprising 18 pairs of 2-letter randomization codes, with each pair allocated to a single school identifier (e.g. School 1 was allocated codes ‘AA’ and AB’, School 2 was allocated codes ‘AC’ and AD’). One 2-letter randomization code within each pair was then randomly assigned to the vitamin D arm of the trial, and the other was assigned to the placebo arm, using a computer-generated random sequence (e.g. ‘AA’ was assigned to ‘vitamin D’, ‘AB’ was assigned to placebo, ‘AC’ was assigned to placebo, ‘AD’ was assigned to ‘vitamin D’). Dr Khudyakov also prepared eighteen separate participant randomization lists (one for each participating school). Each of these participant randomization comprised 999 5-digit numbers, each consisting of a 2-digit school identifier from 01 to 18 that was constant for, and unique to, each list, followed by a 3-digit participant identifier from 001 to 999 (e.g. 01-001, 01-002, 02-001, 02-002). These 5-digit numbers were then randomly assigned to one or other of the 2-letter randomization codes allocated to that school in blocks of ten, using a computer-generated random sequence (e.g. for School 1, sequence was 01-001-AA, 01-002-AB, etc.; for School 2, sequence was 02-001-AC, 02-002-AD, etc.).

Active and placebo capsules were shipped to Mongolia in boxes that were labelled ‘vitamin D’ or ‘placebo’ according to their contents. On arrival, these capsules were packed into bottles, each containing either 1,000 vitamin D capsules or 1,000 placebo capsules. The school randomization list was then used to label these bottles with 2-letter randomization codes according to their contents (i.e. bottles containing vitamin D capsules were labelled with ‘AA’, ‘AD’ or another 2-letter code assigned to the vitamin D arm of the trial, while bottles containing placebo capsules were labelled ‘AB’, ‘AC’ or another 2-letter code assigned to the placebo arm of the trial). Bottling and labelling was performed by Mr Kevin Zinchuk (Brigham and Women’s Hospital, Boston MA), Dr Sabri Bromage (Harvard TH Chan School of Public Health, Boston MA), and Dr Davaasambuu Enkhmaa (National Center for Maternal and Child Health, Ulaanbaatar, Mongolia); none of these individuals was involved with data collection. Children screened at each school were assigned consecutive 5-digit numbers at enrolment by study field workers, and if they were subsequently found to be eligible for randomization (i.e. if their baseline QFT result was negative) then the participant randomization list for their school of attendance was used to determine their allocation, i.e. they received study medication from bottles labelled with the 2-letter code linked to their 5-digit ID in the participant randomization list for the duration of the trial. For example, participant 01-001 would receive study medication labelled ‘AA’ (i.e. vitamin D), 01-002 would receive medication labelled ‘AB’ (i.e. placebo), 02-001 would receive medication labelled ‘AC’ (placebo), and 02-002 would receive medication labelled ‘AD’ (vitamin D) throughout the trial. Copies of the school randomization list were held by members of the DSMB, Dr Khudyakov and Mr Zinchuk. Neither participants nor trial staff had access to it, and treatment allocation was concealed from participants, care providers and all trial staff (including senior investigators and those assessing outcomes) so that the double-blind was maintained. The school randomization list was emailed to ARM and XZ following completion of the trial, who used it to un-blind allocation and to analyse the trial: they were not therefore masked to group assignment during statistical analysis.

### **Table S1.** Fracture incidence by anatomical site, overall and by allocation

|  | Number of participants reporting ≥1 fracture | | | Number of fractures reported | | |
| --- | --- | --- | --- | --- | --- | --- |
|  | Overall | Vitamin D arm | Placebo arm | Overall | Vitamin D arm | Placebo arm |
| Upper limb | 374 | 196 | 178 | 461 | 244 | 217 |
| Lower limb | 134 | 66 | 68 | 163 | 84 | 79 |
| Other | 40 | 24 | 16 | 53 | 30 | 23 |
| Any site | 521 | 268 | 253 | 677 | 358 | 319 |

### **Table S2.** Proportion of participants reporting one or more bone fractures of different types, by allocation

|  |  | **Vitamin D arm (n=4176)** | **Placebo arm (n=4172)** | **Adjusted risk ratio (95% CI) ^(1)^** | **P** |
| --- | --- | --- | --- | --- | --- |
| Confirmation / treatment | Any fracture | 268 (6.4%) | 253 (6.1%) | 1.10 (0.93 to 1.29) | 0.27 |
|  | X-ray confirmed fracture | 259 (6.2%) | 247 (5.9%) | 1.09 (0.93 to 1.29) | 0.29 |
|  | Plaster cast-treated fracture | 238 (5.7%) | 233 (5.6%) | 1.06 (0.89 to 1.26) | 0.50 |
| Fracture site | Upper limb fracture | 196 (4.7%) | 178 (4.3%) | 1.12 (0.92 to 1.35) | 0.27 |
|  | Lower limb fracture | 66 (1.6%) | 68 (1.6%) | 1.15 (0.82 to 1.61) | 0.43 |
|  | Other fracture | 24 (0.6%) | 16 (0.4%) | 1.90 (0.96 to 3.76) | 0.07 |
| Associated trauma | Low-trauma fracture^(2)^ | 104 (2.5%) | 106 (2.5%) | 1.01 (0.79 to 1.30) | 0.93 |
|  | Medium-trauma fracture^(3)^ | 136 (3.3%) | 137 (3.3%) | 1.00 (0.80 to 1.26) | 0.99 |
|  | High-trauma fracture^(4)^ | 24 (0.6%) | 18 (0.4%) | 1.81 (0.92 to 3.54) | 0.09 |

**Abbreviation:** CI, confidence interval.

**Footnotes.** 1, adjusted for random effect of school. 2, defined as ‘slight trauma, such as fall from < 0.5m, e.g. from standing, chair or bed’. 3, defined as ‘moderate trauma, such as fall from 0.5-3m, (e.g. fall down stairs, from a bicycle, roller blades, skateboard or swing); playground scuffles; sport injury’. 4, defined as ‘severe trauma, such as falling from a height >3 metres (e.g. falls from windows or roofs), motor vehicle or pedestrian accident, injury caused by heavy moving or falling objects (e.g., bricks or stones)’.

### **Table S3.** Mean radial speed of sound at 1-, 2- and 3-year follow-up by allocation: overall and by sub-group

|  |  |  | **Vitamin D arm: mean value (s.d.) [n]** | **Placebo arm: mean value (s.d.) [n]** | **Adjusted mean difference (95% CI) ^(1)^** | **P for timepoint** | **Overall P ^(2)^** | **P for interaction** |
| --- | --- | --- | --- | --- | --- | --- | --- | --- |
| Overall |  | 1 year | 3623.8 (136.1) [668] | 3614.3 (126.7) [680] | 8.3 (-5.6 to 22.3) | 0.24 | 0.29 | -- |
|  |  | 2 years | 3612.2 (144.2) [629] | 3618.2 (130.3) [649] | -4.9 (-19.1 to 9.3) | 0.50 |  |  |
|  |  | 3 years | 3645.6 (143.2) [671] | 3646.9 (139.0) [686] | -3.1 (-17.0 to 10.8) | 0.66 |  |  |
| By sex | Male | 1 year | 3618.1 (137.8) [339] | 3608.2 (131.2) [353] | 7.9 (-11.1 to 26.8) | 0.41 | 0.48 | 0.52 |
|  |  | 2 years | 3603.5 (135.4) [322] | 3600.4 (125.4) [332] | 1.8 (-17.6 to 21.1) | 0.86 |  |  |
|  |  | 3 years | 3607.2 (132.4) [336] | 3610.4 (124.4) [358] | -6.9 (-25.9 to 12.0) | 0.47 |  |  |
|  | Female | 1 year | 3629.7 (134.4) [329] | 3621 (121.5) [327] | 7.5 (-12.5 to 27.5) | 0.46 | 0.37 |  |
|  |  | 2 years | 3621.3 (152.6) [307] | 3636.9 (132.9) [317] | -13.5 (-33.9 to 6.9) | 0.19 |  |  |
|  |  | 3 years | 3684.2 (143.4) [335] | 3686.7 (143.3) [328] | -3.1 (-23.1 to 16.9) | 0.76 |  |  |
| By baseline 25(OH)D concentration | <10 ng/ml | 1 year | 3625.7 (134. 5) [233] | 3622.6 (119.2) [254] | 2.1 (-21.0 to 25.2) | 0.86 | 0.79 | 0.79 |
|  |  | 2 years | 3619.7 (144.4) [217] | 3621.6 (139.2) [241] | 1.1 (-22.6 to 24.8) | 0.93 |  |  |
|  |  | 3 years | 3650.4 (148.6) [232] | 3655.9 (141.0) [255] | -5.6 (-28.7 to 17.5) | 0.64 |  |  |
|  | ≥10 ng/ml | 1 year | 3622.8 (137.2) [435] | 3609.4 (130.8) [426] | 14.0 (-3.3 to 31.3) | 0.11 | 0.25 |  |
|  |  | 2 years | 3608.2 (144.1) [412] | 3616.2 (124.9) [408] | -6.1 (-23.7 to 11.5) | 0.50 |  |  |
|  |  | 3 years | 3643.1 (140.4) [439] | 3641.6 (137.7) [431] | 0.4 (-16. 9 to 17.7) | 0.97 |  |  |
| By calcium intake | <500 mg/day | 1 year | 3626.4 (137.2) [423] | 3611.5 (128.8) [409] | 14.0 (-4.0 to 32.0) | 0.13 | 0.29 | 0.18 |
|  |  | 2 years | 3617.5 (138.0) [406] | 3616.7 (130.7) [401] | 2.1 (-16.2 to 20.3) | 0.83 |  |  |
|  |  | 3 years | 3646.5 (141.8) [441] | 3641.4 (142.1) [431] | 4.1 (-13.6 to 21.9) | 0.65 |  |  |
|  | ≥500 mg/day | 1 year | 3615. 8 (133.3) [223] | 3616.5 (121.6) [237] | 1.6 (-22.2 to 25.3) | 0.90 | 0.79 |  |
|  |  | 2 years | 3604.2 (157.0) [213] | 3619.2 (130.5) [235] | -10. 6 (-34.5 to 13.4) | 0.39 |  |  |
|  |  | 3 years | 3645.1 (145.0) [222] | 3655.4 (134.4) [245] | -8.8 (-32.4 to 14.9) | 0.47 |  |  |

### **Figure S1.** Fracture Questions, Case Report Form

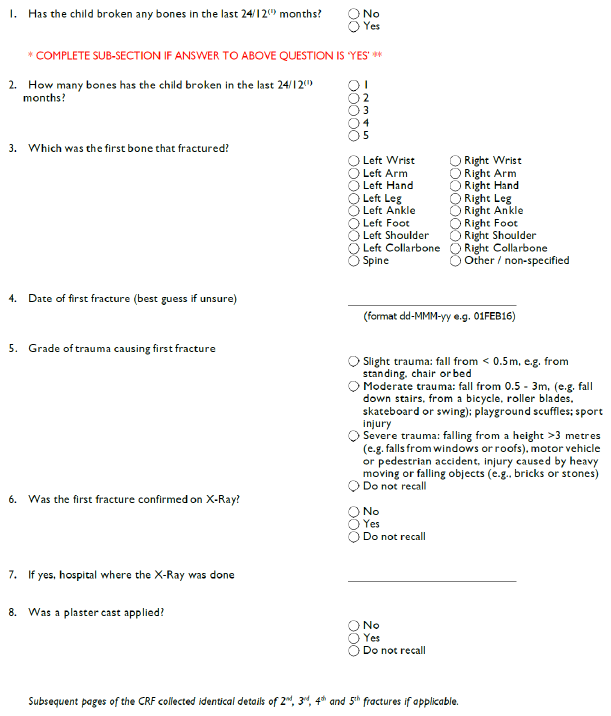

**Footnote:** 1, fractures in previous 24 months were captured at 2-year follow-up, and fractures in previous 12 months were captured at 3-year follow-up.
